# Seed-based morphometry of nodes in the default mode network among patients with Alzheimer’s disease in Klang Valley, Malaysia

**DOI:** 10.1101/2023.08.29.23294758

**Authors:** Nur Hafizah Mohad Azmi, Subapriya Suppiah, Nur Shahidatul Nabila Ibrahim, Buhari Ibrahim, Vengkhata Priya Seriramulu, Malzyfarina Mohamad, Thilakavathi Karuppiah, Nur Farhayu Omar, Normala Ibrahim, Rizzah Mazzuin Razali, Noor Harzana Harrun, Hakimah Mohammad Sallehuddin, Nisha Syed Nasser, Umar Ahmad

**Affiliations:** Department of Radiology, Faculty of Medicine and Health Sciences, Universiti Putra Malaysia, Persiaran Universiti 1, 43400 Serdang, Selangor, Malaysia; Pusat Pengimejan Diagnostik Nuklear, Universiti Putra Malaysia, Persiaran Universiti 1, 43400 Serdang, Selangor, Malaysia; Hospital Sultan Abdul Aziz Shah, Universiti Putra Malaysia, Persiaran UPM-MARDI, 43400 Serdang, Selangor, Malaysia; Malaysian Research Institute on Ageing, Universiti Putra Malaysia, Selangor, Malaysia; Department of Physiology, Faculty of Basic Medical Sciences, Bauchi State University PMB 65, Gadau, Nigeria; Faculty of Health Sciences, Universiti Kebangsaan Malaysia, Jalan Raja Muda Abdul Aziz, 50300 Kuala Lumpur, Malaysia; Department of Biomedical Sciences, Faculty of Medicine and Health Sciences, Universiti Putra Malaysia, Persiaran Universiti 1, 43400 Serdang, Selangor, Malaysia; Department of Psychiatry, Faculty of Medicine and Health Sciences, Universiti Putra Malaysia, Persiaran Universiti 1, 43400 Serdang, Selangor, Malaysia; Department of Medicine, Hospital Kuala Lumpur, Jalan Pahang, 50300 Kuala Lumpur, Malaysia; Klinik Kesihatan Pandamaran, Persiaran Raja Muda Musa, 4200 Klang, Selangor, Malaysia; Department of Medicine, Faculty of Medicine and Health Sciences, Universiti Putra Malaysia, Persiaran Universiti 1, 43400 Serdang, Selangor, Malaysia; Nanyang Technological University, 50 Nanyang Avenue, 639798, Singapore; Molecular Genetics Informatics, Department of Anatomy, Faculty of Basic Medical Science, Bauchi State University, Gadau, Bauchi, PMB 65, Nigeria; Institute of Pathogen Genomics, Centre for Laboratory and Systems Networks, Africa Centres for Disease Control and Prevention (Africa CDC), African Union Commission, P.O. Box 3243, Addis Ababa, Ethiopia

## Abstract

The default mode network (DMN) is a prominent neural network in the human brain that exhibits a substantial association with Alzheimer’s disease (AD). Functional connectivity (FC) and grey matter volume (GMV) were reported to differ between AD and healthy controls (HC). Nevertheless, available evidence is scarce regarding the structural and functional alterations observed in individuals diagnosed with Alzheimer’s disease (AD) within the context of Malaysia. A prospective cross-sectional study was conducted in the Klang Valley region of Malaysia. A total of 22 participants were enlisted for the study, following a thorough clinical assessment completed by geriatricians. The participants underwent a series of neuropsychological tests, including the Montreal Cognitive Assessment (MoCA), Mini-Mental State Examination (MMSE), and Clinical Dementia Rating (CDR). The participants were classified into two groups, namely AD (Alzheimer’s disease) and HC (healthy controls), before the acquisition of resting-state functional magnetic resonance imaging (Rs-fMRI) images. The analysis of voxel-based morphometry (VBM) was conducted using SPM 12, a widely used software package in the field of neuroimaging, implemented in MATLAB. The primary objective of this analysis was to assess the grey matter volume (GMV). The CONN toolbox was employed to assess the functional connectivity (FC) and activation patterns of the nodes inside the default mode network (DMN). In this pilot project, a cohort of 22 participants was enlisted, consisting of 11 individuals with Alzheimer’s disease (AD) with an age range of 64-84 years (mean age 76.36 ± 0.52) and 11 healthy controls (HC) with an age range of 64-79 years (mean age 69.91 ± 5.34). In the Alzheimer’s disease (AD) group, there was a reduction in grey matter volume (GMV) observed in several brain regions when compared to the healthy control (HC) group. Specifically, decreased GMV was found in the right and left inferior temporal gyrus, left superior frontal gyrus, right superior frontal gyrus medial segment, right gyrus rectus, right temporal lobe, left putamen, and right precuneus, respectively. The significance level for the Rs-FC analysis was established at a cluster-size corrected p-value of less than 0.05. A notable reduction in the activation of the nodes within the default mode network (DMN) was observed in individuals with Alzheimer’s disease (AD) compared to healthy controls (HC). This drop was notably evident in the functional connectivity of the precuneus and anterior cingulate cortex in both AD and HC groups, as well as in the comparison between AD and HC groups. Resting-state functional magnetic resonance imaging (fMRI) can identify specific imaging biomarkers associated with Alzheimer’s disease by analysing grey matter volume (GMV) and default mode network (DMN) functional connectivity (FC) profiles. Consequently, there is promise for utilising resting-state fMRI as a non-invasive approach to enhance the detection and diagnosis of Alzheimer’s disease within the Malaysian community.

## 1.0 INTRODUCTION

Alzheimer’s disease (AD) is the most prevalent form of dementia and a progressive neurodegenerative disorder associated with memory loss. Historically, AD begins with atrophy in the hippocampus and spreads to other brain regions due to accelerated neuronal inflammation and death(1, 2). AD is one of the important illnesses of the elderly, in which the incidence has increased significantly in recent years (3). An earlier study among senior Malaysian citizens revealed that older age, no formal education, female gender, low self-rated health quality, and Malay or Bumiputera ethnicity were significant risk factors for dementia (4, 5). Moreover, the estimated number of people with dementia in Malaysia reported by Alzheimer’s Disease International in 2030 is 261,000 (6), 0.72% of the estimated population of 36.09 million, according to the UN World Population Prospects 2019.

Currently, neurophysiological tests, such as the Montreal Cognitive Assessment (MoCA), Mini-Mental State Examination (MMSE), and Clinical Dementia Rating (CDR) Scores, are routinely used to diagnose AD (7–9). The MoCA should not be used as a substitute for a more in-depth neuropsychological assessment (10–12). Meanwhile, the MMSE is an effective instrument for screening dementia in older persons with basic literacy abilities. Nonetheless, it has a high risk of misclassification in illiterate older persons, which has significant implications for detecting AD in developing nations with low literacy rates among older adults (13). The CDR is a global dementia rating scale that determines the presence of dementia and evaluates cognitive change (14). Consequently, neuroimaging research has been utilised to support the diagnosis of Alzheimer’s disease (15, 16).

Neuroimaging studies using T1-weighted MRI images were used to evaluate abnormalities in brain structure (17–19). Structural MRI images were processed using voxel-based morphometry (VBM). Subsequently, the grey matter volume (GMV) loss in brain structures was identified in AD patients, which usually occurred first in the temporoparietal lobes (20–23). VBM, an MRI analytic technique, can be used to investigate non-invasively the morphological abnormalities of the brain (17, 24). VBM is a computational tool for examining anatomical sections of the neuronal cortices and quantifying differences in local brain tissue concentrations, mainly in the grey matter (25, 26).

Grey matter atrophy was specific to an increased risk for AD (27). Grey matter density (GMD) at each voxel can be compared across the brain between AD patients (28, 29). The increased GMD that was inversely correlated with decreased GMV was found involving the medial temporal lobe extending to areas in the temporal gyri, precuneus, insular and cingulate cortex, and caudate nucleus in AD (29, 30).

Meanwhile, in rs-fMRI neuroimaging studies the seed-based analysis (SBA) method was utilised to analyse functional connectivity (FC) based on blood oxygen level-dependent sequence from rs-fMRI examinations (31, 32). SBA is useful for the detailed analysis of a particular Region of Interest (ROI), to measure functional changes between participants and to reveal the FC among the nodes of the DMN (19, 33). Historically, DMN FC has been implicated with altered brain morphometry in various neurological and psychological disorders (34). Previous study has revealed that there is decreased FC that is widespread in the brain of AD compared to HC participants, particularly involving the nodes of the DMN (35). In AD participants, the MN FC was noted to be decreased in certain brain areas that were significantly correlated with reduced cortical thickness, namely in the superior temporal, supramarginal gyrus of the left cerebral hemisphere (36, 37).

DMN has been implicated in numerous studies involving FC in AD participants. Based on previous fMRI studies, there is *a priori* knowledge regarding the affected nodes of the DMN, which include the precuneus (Prec), posterior cingulate cortex (PCC), retro-splenial cortex, medial parietal cortex (MPC), lateral parietal cortex (LPC), and inferior parietal cortex (IPC), medial prefrontal cortex (mPFC), and medial temporal gyrus (MTG)(4, 36, 38). Despite many studies conducted in Caucasian and North Asian populations, there is a lack of data regarding rs-fMRI in Malaysia to elucidate the changes that occur in AD and identify imaging biomarkers in our population.

We hypothesise that there will be a significant difference in the neuropsychological profile of AD compared with HC. We also hypothesise there will be altered GMV in *a priori* areas of the brain of AD participants. Additionally, we hypothesise that the severity of AD can be correlated with marked atrophy of GMV and a significant reduction in rs-fMRI neuronal FC.

We aimed to describe the processing of structural MRI data using VBM that can help to detect the differences in regional GMV in AD patients compared to HC. Another objective of this study was to identify the differences in rs-fMRI FC in AD compared to HC.

## 2.0 MATERIALS AND METHOD

### 2.1 Study design and subject recruitment

This is a prospective cross-sectional study that received ethical clearance from the Universiti Putra Malaysia ethical committee with ethical clearance number JKEUPM-2019-328 and Malaysian national ethical clearance, MREC (NMRR-19-2719-49105). The data for the study was collected from March 2021 to June 2022. The database of AD patients attending Hospital Kuala Lumpur (HKL) memory clinic, Klinik Kesihatan Pandamaran Klang, and Hospital Sultan Abdul Aziz Shah Universiti Putra Malaysia (HSAAS UPM) were surveyed to recruit suitable AD participants. We recruited age-matched cognitively healthy controls (HC) by sending out flyers to the community and posting them on community bulletin boards. The participants who met the inclusion criteria were selected for this study. Following the principles outlined in the Declaration of Helsinki 1964, participation in the study was voluntary, and informed consent was acquired from potential participants before recruitment. The participants were compensated, and all data were anonymised.

### 2.2 Inclusion and exclusion criteria

The inclusion criteria for participants with a clinically verified diagnosis of AD as well as being Malaysian citizens, aged between 55 and 90 years old. The physicians classified the participants into AD and HC groups using DSM-5 and MoCA, MMSE, and CDR scores based on their clinical assessment. Participants in the HC group were required to have a good memory and no brain diseases, including cancer and stroke. The participants did not suffer from claustrophobia, had no metal implant, and cooperated for the rs-fMRI scan. Exclusion criteria are non-citizens of Malaysia and those with neurological diseases other than AD. Relative and absolute contraindications for MRI examination include claustrophobia, irremovable metallic implants that are not MRI compatible, and electronic implants, such as pacemakers, cochlear or ear implants, and metallic tattoos.

### 2.3 Alzheimer’s disease and neurophysiological assessment

The participants were administered with several questionnaires, which included the MoCA, MMSE and CDR scores.

### 2.4 Montreal Cognitive Assessment (MoCA)

An 8-item self-reported MoCA questionnaire was used. evaluate short-term memory, executive functions, visuospatial abilities, attention and concentration including working memory, as well as language and orientation to time and place). The scores range are 5 points for a short-term memory recall task involving two learning trials of five nouns and delayed recall after approximately five minutes, for visuospatial abilities using 3 points for clock-drawing and 1 point for a three-dimensional cube copy. A verbal language task was also administered. One point was given for attention, concentration, and working memory, which were assessed using a sustained attention task; then 3 points for a serial numbers’ subtraction task, and 1-point digits span forward and digit span backwards task. Three points for language are assessed using a three-item confrontation naming task with low-familiarity animals (lion, camel, rhinoceros), 2 points for repetition of two syntactically complex sentences, and the fluency above task. Finally, 6 points for orientation to time and place are evaluated by asking the subject for the date and the city where the test occurred. The MoCA score ranges from 0 to 30. After evaluating the MoCA questionnaire, a normal score is regarded as 26 or higher. People without cognitive impairment scored an average of 27.4 in research, while those with mild cognitive impairment (MCI) scored 22.1 and those with Alzheimer’s disease scored 16.2 (39, 40).

### 2.5 Screening tool: The Mini-Mental State Examination (MMSE)

A five-item self-reported MMSE or Folstein test is a 30-point questionnaire. The questionnaire was used to screen for dementia. The scores ranged from 10 points for orientation (time and place), 3 points for registration. 5 points for Attention and calculation, 3 points for recall and 9 points for language (language, repetition, and complex commands). After evaluating the MMSE questionnaire, participants who scored <26 for Alzheimer’s disease and those who scored ≥26 were normal (41)

### 2.6 Clinical Dementia Rating (CDR)

The CDR tool is a numerical scale that is used worldwide to identify the dementia severity stages by assessing dementia symptoms (42). A qualified medical or psychological personnel will determine a patient’s cognitive and functional performance in 6 cognitive areas: orientation, memory, judgement & solving problems, home & hobbies, community affairs, and personal care, by administering a structured interview-based protocol developed in 1982 by Charles Hughes (43). The results of each of these are added together to get a composite score that ranges from 0 to 3. This score is useful for characterising and tracking a patient’s level of cognitive impairment/dementia: 0 = Normal, 0.5 = Questionable Dementia, 1 = Mild Dementia, 2 = Moderate Dementia, and 3 = Severe Dementia.

### 2.7 MRI data Acquisition

The MRI was performed on a Siemens 3.0T scanner (PRISMA, Siemens, Erlangen, Germany). A 12-channel head coil was used for structural MRI. T1 MPRAGE MRI data with high resolution was acquired. The sequence’s parameters were as follows: TR = 2300ms, TE = 2.27ms, TI=1100ms, number of slices =160, ascending sagittal oriented, FOV =256 x 256 mm2, matrix size =256 x 256, and slice thickness=1mm.

Blood-oxygen-level-dependent (BOLD) Imaging was performed using an echo-planar imaging (EPI) sequence. The rs-fMRI images were obtained with a field of strength of 3.0 Tesla, a repetition time of 2500msec, an echo time of 30ms, a flip angle of 90°, matrix 64 x 64, 250 volumes, 38 slices per volume, and a slice thickness of 3.5mm. The voxel size: is 2.5 x 2.5 x 3 mm^3^. The phase encoding direction was from anterior to posterior, with a. Participants were asked to lie down with closed eyes but not fall asleep.

### 2.8 Pre-processing Voxel-Based Morphometry (VBM) Analysis

The VBM toolbox in the Statistical Parametric Mapping software (SPM 12, http://www.fil.ion.ucl.ac.uk/spm/software/spm12) implemented in MATLAB was used to pre- process structural images (2, 44, 45).

First, all images were checked for artefacts and structural abnormalities. Secondly, temporal processing involved slice timing, and thirdly, spatial processing involved realignment and estimate, set origin, co-registration, normalisation, and smoothing were performed. The group-specific AD and HC templates were utilized to reduce variability among participants. The Asian brain map template was then used to normalise the images using the “DARTEL Normalize to Montreal Neurological Institute MNI Space” program. The volume for a specific ROI based on a priori knowledge was generated using T1-weighted images that were spatially registered to the MNI template. Based on spatial registration and modulated images of the grey matter that mirrored the tissue volumes, segmented images of the GMD and GMV were retained to measure the number of volume changes. After that, a Gaussian filter was used to smooth the normalised brain pictures (8mm FWHM). The family-wise error (FWE) was used for multiple comparisons, using a p<0.05 threshold. The threshold in the SPM analysis, which was deemed uncorrected for FWE, was decreased to p<0.001 to find regions with low signals.

The GMV differences between the AD and HC groups were evaluated using 2-sample t-tests in SPM12. A threshold of voxel-wise uncorrected p < 0.001 and cluster-level p < 0.05 FWE correction were applied to the rs-fMRI data. Uncorrected p<0.001 was used due to the small sample size.

### 2.9 Resting-state functional Connectivity (Rs-FC) analysis using seed-based analysis

Rs-FC analysis was performed using a seed-based approach using the CONN toolbox v20.b (http://www.nitrc.org/projects/conn). We conducted whole-brain research, followed by seed-based analysis (SBA) using ROIs based on *a priori* knowledge. The functional images were pre- processed with SPM12 software by applying the following steps: slice-timing correction; spatial realignment; co-registration to the T1-weighted anatomical image; spatial normalization to the MNI space, and smoothing (44). The significance level was set at p<0.001, FWE uncorrected.

### 2.10 Statistical Analysis

SPM 12 and Statistical Package for the Social Sciences (SPSS software Version 25.0, SPSS Inc., Chicago, IL, USA) was used for statistical analysis. The descriptive statistic was performed to analyse participants’ socio-demographic data, and the chi-square test was used to analyse the association between two groups with neuropsychological tests. Two sample t-tests were used to compare the differences in brain GMV in AD versus HC participants using VBM. The significance level was set at a p-value < 0.05. Regression analysis was performed in ROI-based DMN data analysis to identify brain activation regions in AD participants using SBA of rs-fMRI data. The significance level was set at a voxel threshold p-value < 0.001 and cluster threshold p-value < 0.05.

## 3.0 RESULTS

### 3.1 Demographic characteristics

Twenty-two participants were recruited for this study. The participants were divided into AD and HC groups. Both groups were compared using age distribution, gender, education level and marital status, and neuropsychological tests i.e., MoCA, MMSE and CDR (Table 1). Thus, there were eleven participants in each group, i.e., Alzheimer’s disease (AD) and healthy control (HC) groups as shown in **Table 1**. The average age of the AD and HC groups is 64-84(76.36 ±0.52) and 6-79(69.91 ±5.34) years old respectively. An independent-sample t-test indicated that age was significantly different between AD and HC t (20) =-2.66, *p*=0.015. Therefore, a significant association between AD and HC was found in **Table 1**.

**Table 1:**
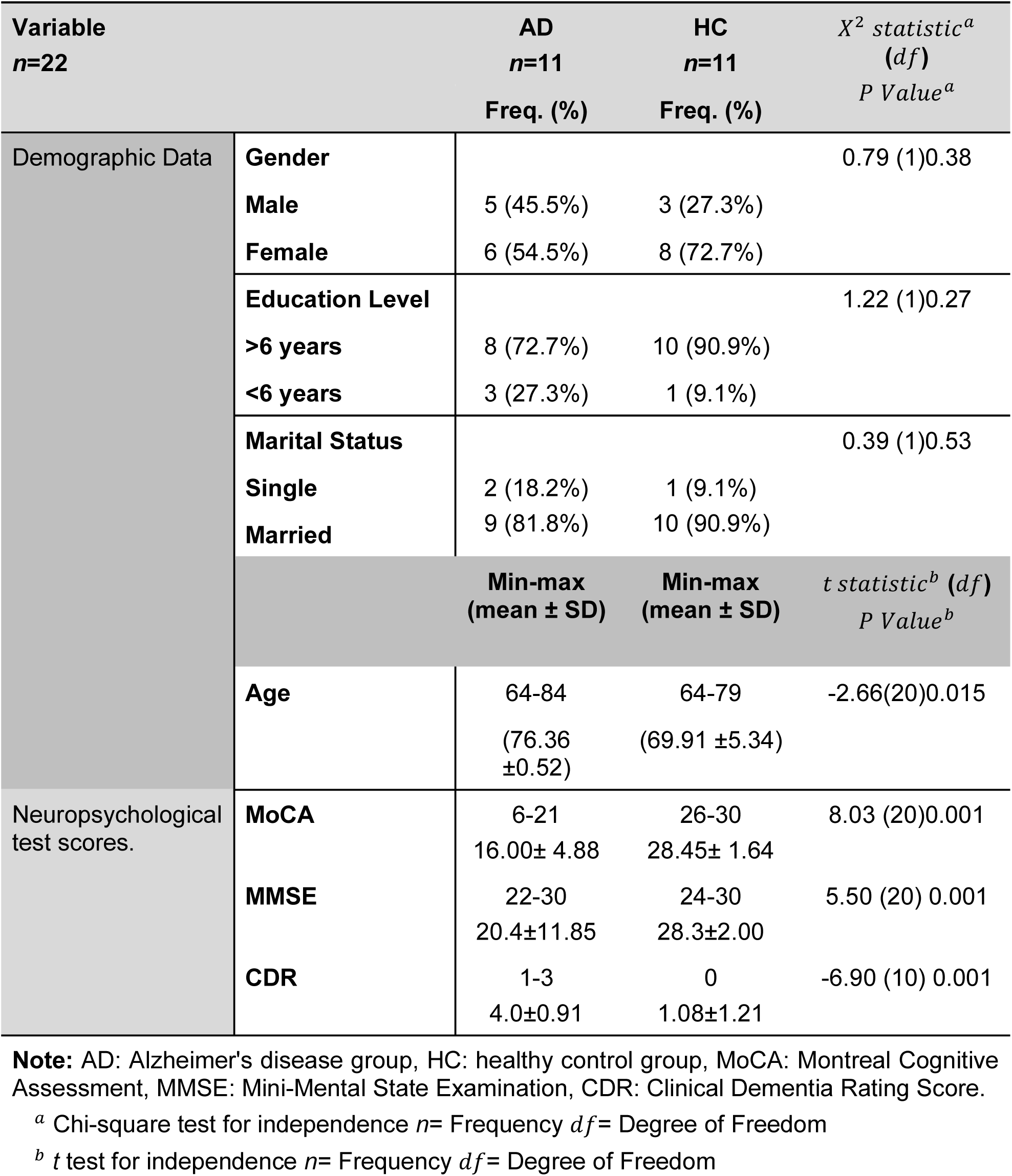
Comparison of sociodemographic and neuropsychological profile of AD with HC.

An independent-sample t-test indicated that MoCA, MMSE and CDR were significantly different between AD and HC t (20) =8.03, *P*=0.001, t (20) =5.50, *P*=0.001 and t (10) =-6.90, *P*=0.001. Therefore, a significant association between AD and HC is referred to in **Table 1**.

A Chi-square test for independence indicated that the AD and HC participants between male and female, < six years education level and >six years education level, single and marital status are not significantly different (gender; *P*=0.38 education level; P=0.27, and marital status; *P*=0.53 respectively). Therefore, there is no significant association between gender, education level and marital status with AD and HC refer to **Table 1**.

### 3.2 Neuropsychological assessment test

The descriptive statistics of neuropsychological test scores for the AD and HC group with neurophysiological assessment parameters are tabulated **(see Table 1)**. In MoCA HC groups scores 26-30, which indicates Normal, and AD group scores 6-21, which indicates Normal. Using MMSE, we detected that the HC group had scores ranging from 24-30 in keeping with no cognitive impairment, and the AD group scores 22-30, ranging from mild to severe cognitive impairment, respectively. While in CDR n=22, there were participants in the HC group detected with normal daily functioning and in the AD, group were detected with mild dementia and impairment of daily living activities.

### 3.3 Voxel-based morphometry analysis

Right inferior temporal gyrus (ITG r), left inferior temporal gyrus (ITG l), left superior frontal gyrus (SFG l), right superior frontal gyrus medial segment (MSFG r), right gyrus rectus or straight gyrus, right temporal lobe, left putamen and right precuneus was found to be high grey matter density or voxel density for AD compared to the HC group (p<0.001, FWE corrected) as shown in **Table 2** and **Figure 1**.

**Figure 1.**
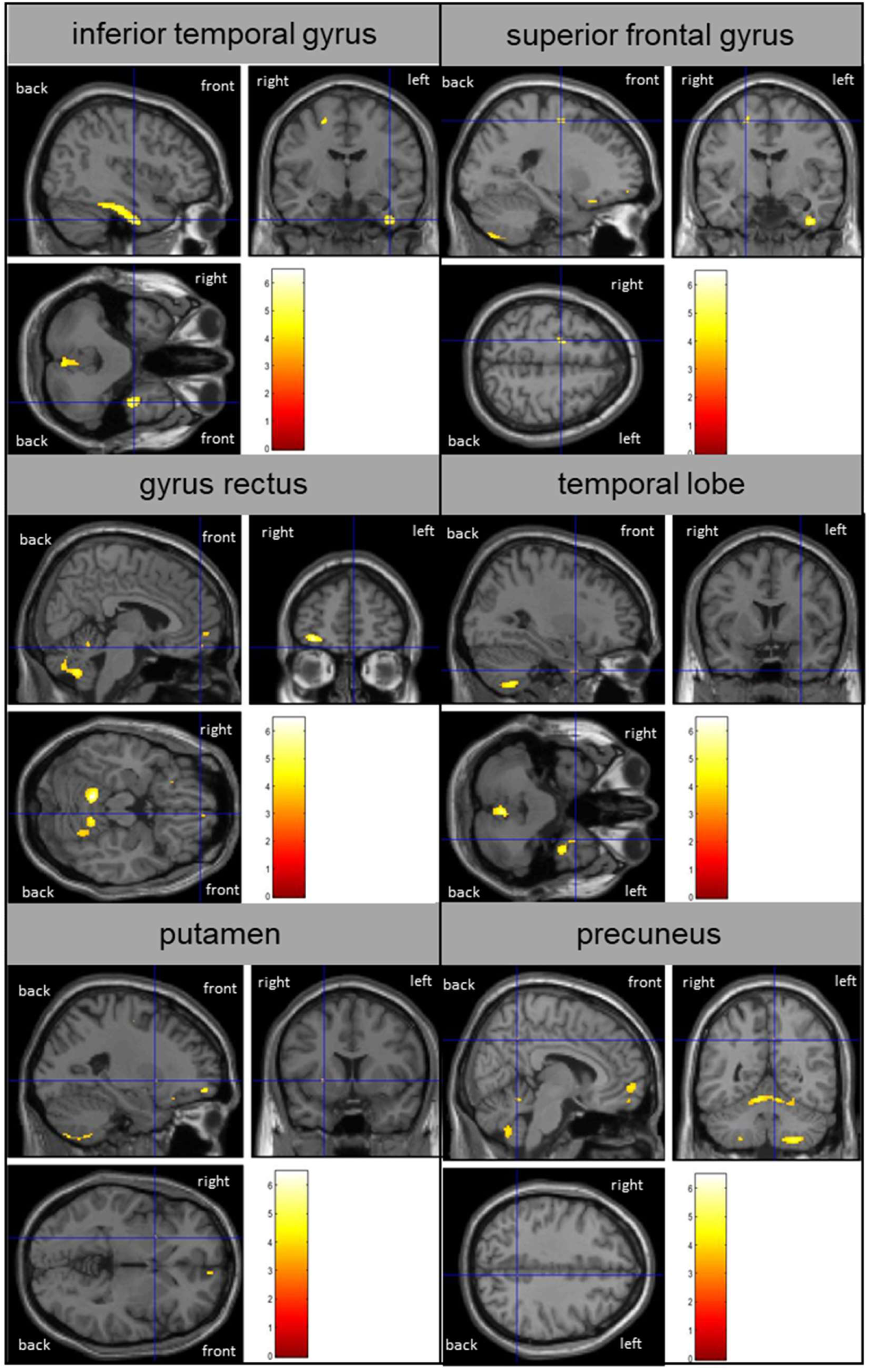
VBM results had different density and reduced grey matter volume in AD more than HC participants using T1 MRI structural data, corrected p-value 0.001 images (p<0.05, FWE corrected)

**Table 2:** Tabulated values of regional difference in voxel density for AD > HC group and HC > AD corrected p-value 0.001 images (p<0.05, FWE corrected)

| AD>HC |  | Side | Cluster<br>Peak<br>mm mm<br>mm | Voxel | Peak T | P Level<br>(FWE) |
| --- | --- | --- | --- | --- | --- | --- |
| Inferior temporal gyrus | ITG r | Right | 40 -9 -38 | 849 | 5.57 | <0.001 |
| Inferior temporal gyrus | ITG l | Left | -45 -32 -27 | 17 | 3.73 | <0.001 |
| Superior frontal gyrus | SFG l | Left | -21 -8 56 | 55 | 4.91 | <0.001 |
| Superior frontal gyrus<br>medial segment | MSFG<br>r | Right | 8 58 -4 | 57 | 4.25 | <0.001 |
| gyrus rectus |  | Right | 6 52 -18 | 7 | 3.67 | <0.001 |
| temporal lobe |  | Right | 30 6 -40 | 11 | 3.66 | <0.001 |
| Putamen |  | Left | -24 9 -2 | 5 | 3.15 | <0.001 |
| Precuneus |  | Right | 8 -52 39 | 2 | 3.57 | <0.001 |
| AD<HC |  | Side | Cluster<br>Peak | Voxel | Peak T | P Level<br>(FWE) |
| - |  | - | - | - | - | - |

**Note:** AD: Alzheimer's disease group, HC: healthy control group

### 3.4 Seed-based functional connectivity analysis

**Table 3** and **Figure 2** show the brain regions with significant functional connectivity differences within the seed between AD and HC participants. The warm colours represent high values and cool colours represent the opposite. High values were found in the precuneus, and anterior cingulate gyrus (ACG) for both groups. In HC participants, the high values were found in the right and left lateral occipital, and right and left frontal lobes. For AD participants, the high values were found in the superior left occipital cortex and superior lateral occipital cortex. FC found no significant differences between AD and HC.

**Figure 2.**
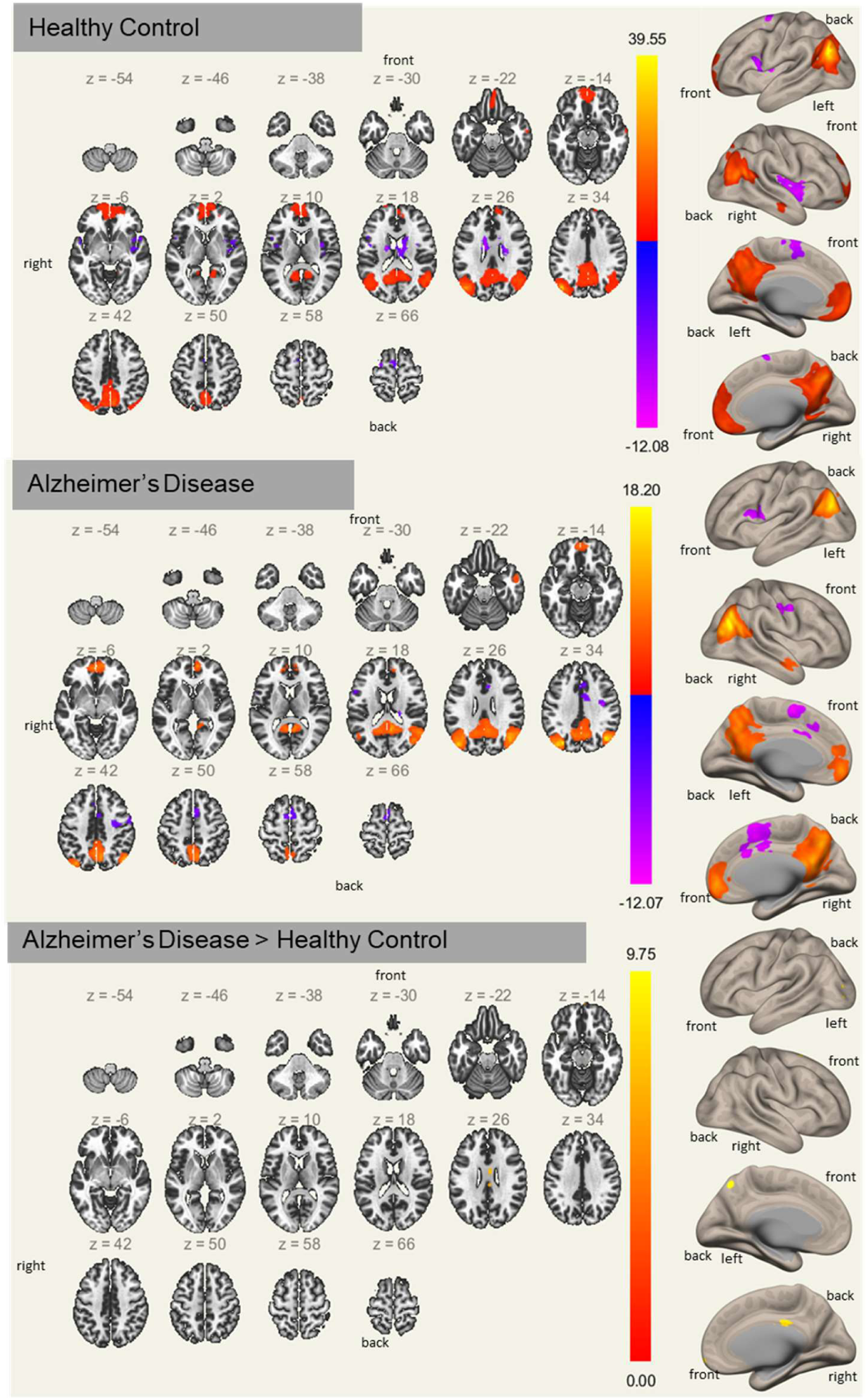
FC Analysis differences between AD and HC. Activation maps are graphical representations of activation regions in the brain. Where hot colours represent greater mean regional activation between specific regions and cold colours represent lower activation group differences. Activation values based on T values (i.e., activation color bar range -12.08 to 39.55 for HC, -12.07 to 18.20 for AD and 0 to 9.75 for differences between AD and HC). FC: Functional connectivity. Rs-FC analysis found the significance level was set at p<0.001, cluster-size corrected p<0.05.

**Table 3:** Tabulated values of regional difference in seed-based Rs-fMRI functional connectivity in AD, HC, AD>HC and HC cluster-size corrected p<0.05.

|  |  | Side | Cluster Size<br>(voxels) | MNI coordinates<br>x y z | P Value |
| --- | --- | --- | --- | --- | --- |
| **HC** |  |  |  |  |  |
| Precuneus |  |  | 4138 | -40 -78 +32 | 0.452 |
| Lateral Occipital Cortex |  | Left | 2957 |  |  |
| Lateral Occipital Cortex |  | Right | 2745 |  |  |
| Frontal Lobe | FP | Right | 1238 | +00 +58 -02 |  |
| Frontal Lobe | FP | Left | 1082 |  |  |
| Cingulate Gyrus | AC | Anterior | 934 |  |  |
| **AD** |  |  |  |  |  |
| Precuneus |  |  | 3755 | +02 -68 +50 | 0.246 |
| Lateral occipital cortex | sLOCI | Superior, Left | 2301 |  |  |
| Cingulate gyrus | AC | Anterior | 1014 | +02 +52 -04 |  |
| Lateral occipital cortex |  | Superior | 1599 | +46 -70 +34 |  |
| **AD>HC** |  |  |  |  |  |
| - |  |  |  |  |  |

**Note:** AD: Alzheimer's disease group, HC: healthy control group

## 4.0 DISCUSSION

This study prospectively recruited AD participants in Klang Valley, Malaysia and conducted neuropsychological tests and fMRI examinations for the participants and age-matched healthy controls. There were significantly reduced MMSE, MoCA and CDR values among the AD participants compared to the controls, which indicates the tools as a good screening test to identify neurocognitive deficits in the participants. The MoCA neuropsychological exam was found to correlate with the MMSE similar to other studies (9). To specify the outcome, we examined the CDR scores and found that the AD participants had performance levels in the mild to severe categories (46). The CDR score also made it easier to identify the participants with AD compared to HC, as well as identify the severity of the neurocognitive deficit based on mild, moderate and severe stages of the disease. Nevertheless, these tests are semi-objective and have a variable level of specificity.

Thus, this is the reason that we conducted neuroimaging studies that can act as a more specific biomarker of AD. fMRI can detect small changes in the GMV and FC of neuronal networks that help to differentiate the features of AD from HC. Based on our rs-MRI study, using morphological data from the structural MRI images and functional data from the rs-fMRI examination, we evaluated the intrinsic neural activity in participants with AD. We identified some similarities with other studies such as atrophy of the precuneus (47). We detected that AD had reduced GMV at the ITG r and ITG l (48), SFG l (49), MSFG r (49), right gyrus rectus or straight gyrus (49) right temporal lobe (50), left putamen (51) and right precuneus. Furthermore, Guo, Chen (52) found decreased GMV in the ventral precuneus and postulated that this region helps to boost the efficiency of conscious processes, allowing individuals to transition between different temporal frames depending on the situation and leading to more balanced time perspectives, which then becomes impaired in AD patients. Interestingly, no brain region had significantly atrophied GMV at any specific node in the age-matched HC compared to the AD participants, as we have hypothesised because of accelerated degeneration that occurs in AD compared to normal ageing (53).

Rs-MRI detected reduced FC in regions of the DMN similar to previous findings by Park, Park (36). In our study, we also detected reduced FC in AD among our Malaysia population, specifically deactivation in the precuneus and the anterior cingulate gyrus. It is hypothesised that reduced activations in the regions of the DMN are caused by accelerated neurodegeneration that occurs in the related nodes, which can be detected at an early stage using fMRI with improved diagnostic accuracy, of approximately 82.6% sensitivity and 79.1% specificity, respectively in discriminating AD from healthy controls (54).

Limitations of our study include a small sample size, which was due to our recruitment period occurring during the COVID-19 pandemic and restricted by the movement control orders causing the participants to have difficulty to have access to the imaging centre. Furthermore, some AD participants were not cooperative with the scan, and the data had to be removed from the final analysis due to artefacts.

Our future recommendation is to incorporate a multicentre study protocol to improve the sample size and also to utilise artificial intelligence algorithms that can extract imaging features in an automated pipeline for improved diagnostic accuracy. In addition, with better sample size and adequate representation of all the stages of AD, future studies can extract specific imaging features that can be utilised as early markers of the disease in the Alzheimer’s disease continuum.

## 5.0 CONCLUSION

Using the VBM method, we identified reduced GMV in the precuneus among participants with Alzheimer’s disease. Our results support our hypothesis that there will be altered GMV and network functional connectivity in brain regions of the brain similar to our *a priori* knowledge.

## Data Availability

All data produced in the present study are available upon reasonable request to the authors.

## Supplementary Materials

## Author Contributions

NHMA, SS, NSNI, and VPS, were involved in data collection and analysis. AAR also performed data interpretation and prepared the first draft of the manuscript. NHMA and SS were responsible for the conceptual framework and study design, secured financial support, conducted data interpretation and supervised the project. MM also helped in formulating the conceptual framework and study design and the data analysis and interpretation. NHMA and SS conducted the literature search, data analysis, and data interpretation. NHMA, SS, TK, FO, and BI were involved in the study design, project supervision, and verification of the analytical methods. YNT was involved in securing part of the financial support for this study and data collection. NHMA, SS, RMR, HNH, HS, NS, and UA were involved in the conceptual framework, verification of analytical methods, and data interpretation. All the authors were involved in editing and verifying the final completed manuscript.

## Acknowledgement

This research was funded by the Fundamental Research Grant Scheme (FRGS 06- 02-14-1497FR/5524581) awarded by the Malaysian Ministry of Higher Education under grant number 5540244. The Ministry of Health Malaysia and the Malaysian Society of Radiographers are also thanked for their unwavering support of this research. We are also grateful to the personnel at Pusat Pengimejan Diagnostik Nuklear, UPM, who contributed directly or indirectly to the data collection.

## Ethical clearance

This study was approved by the Ethics Committee of Research Involving Human Participants of Universiti Putra Malaysia (JKEUPM-2019-328) and MREC (NMRR-19-2719-49105).

